# Longitudinal Enrichment of Eosinophilic Esophagitis in Children with AD: The MPAACH Cohort

**DOI:** 10.1101/2025.09.22.25336085

**Authors:** Wan Chi Chang, Lisa J. Martin, Latha Satish, Lindsey Williams, Mindy Hammonds, Seth Jenkins, Garrett Osswald, Julie Caldwell, Marc E. Rothenberg, Gurjit K. Khurana Hershey, Jocelyn M. Biagini

**Affiliations:** Division of Asthma Research, Cincinnati Children’s Hospital Medical Center, Cincinnati, OH; Division of Human Genetics, Cincinnati Children’s Hospital Medical Center, Cincinnati, OH; Department of Pediatrics, University of Cincinnati College of Medicine, Cincinnati, OH; Division of Allergy and Immunology, Cincinnati Children’s Hospital Medical Center, Cincinnati, OH

**Author notes:** **Corresponding Author:** Jocelyn M Biagini, PhD, Division of Asthma Research, Cincinnati Children’s Hospital Medical Center, 3333 Burnet Avenue, Cincinnati, OH, 45229, USA.

**Keywords:** atopic dermatitis, eosinophilic esophagitis, atopic march, children

## Abstract

**Introduction:** The atopic march refers to the natural history of the sequential development of atopic dermatitis (AD), food allergy (FA), asthma and allergic rhinitis (AR) in childhood, but there is significant heterogeneity in the timing and order of disease presentation. Eosinophilic esophagitis (EoE) is an emerging food-induced inflammatory disease of the esophagus that shares clinical and immunologic features, genetic susceptibility and comorbidities with other march members. We sought to investigate the incidence and prevalence of EoE in a cohort of children with AD and elucidate the individual trajectories of sensitization and allergic comorbidity development.

**Methods:** We evaluated 700 children participating in the Mechanisms of Progression of Atopic Dermatitis to Asthma in Children (MPAACH) cohort. FA was defined as parental report of a physician diagnosis. AR was defined by a positive skin prick test to ≥1 aeroallergen and symptoms. Asthma was defined by pulmonary function testing and symptoms according to ATS criteria. Participants were classified as having EoE if they had ≥15 eosinophils per high-power field in at least one esophageal biopsy.

**Results:** Of the 700 children in MPAACH, 10 developed EoE, all after AD onset. The cumulative incidence of EoE in MPAACH was 0.7% by age 2, 0.7% between ages 2 and 5, and 1.6% between ages 5 and 8. The prevalence of EoE in MPAACH is 1.4% (95%CI 0.5-2.3%) and the median age at EoE diagnosis was 3.7 years (interquartile range (IQR): 1.8-6.5). Despite no difference in skin barrier quality or AD severity, children with EoE were more likely to have food sensitization (60% vs. 28%, p=0.039) and FA (70% vs. 13%, p<0.001). 90% of children with EoE had ≥1 allergic comorbidity compared to 49% of the MPAACH cohort (p=0.009). Of these 9, 6 developed FA and/or AR prior to their EoE diagnosis and 3 children developed FA, AR, and/or asthma after EoE onset.

**Conclusions:** Our data support the inclusion of EoE as a fifth member of the march, with new incidence estimates showing enrichment in children with AD, underscoring early-life AD as a risk factor for EoE. The higher prevalence of food sensitization and FA despite no difference in skin barrier quality or AD severity suggests the esophagus as the site of allergen penetration independent of the skin. These data support screening for EoE symptoms in children with early allergic manifestations.

## Introduction

The atopic march refers to the natural history of allergic disease manifestations as they develop in childhood^1^. Classically, the march begins with atopic dermatitis (AD), followed sequentially by food allergy (FA) asthma and allergic rhinitis (AR)^1^, but there is significant heterogeneity in the timing, order, and organ(s) affected^2^. While AD is strongly associated with the development of FA, asthma and AR, most individuals do not follow the traditional atopic march sequence^2^. A 2014 latent class analysis of 9,801 children from 2 UK birth cohorts identified 8 distinct atopic march trajectory profiles of AD, wheeze and AR^3^. A 2019 NIH workshop concluded that the atopic march needs to be revised to include this heterogeneity and incorporate the pathogenesis of the various combinations of the atopic march. We designed the <u>M</u>echanisms of <u>P</u>rogression of <u>A</u>topic Dermatitis (AD) to <u>A</u>sthma in <u>CH</u>ildren (MPAACH) cohort^4^, the first US longitudinal early life cohort of AD, to meet this need.

Eosinophilic esophagitis (EoE) is an emerging chronic, food-induced, allergic inflammatory disease characterized by marked esophageal eosinophilia and esophageal dysfunction^5^. There is evidence that EoE could be considered a fifth member of the atopic march given it shares clinical features, immunologic pathways, genetic susceptibility loci, and comorbidities with other atopic march members^5^. Despite these observations, rigorous epidemiologic studies of EoE in the context of the allergic march are limited, largely because EoE is relatively rare (0.14% in the general population)^6^. Since FA, AR and asthma are enriched in children with AD, we sought to investigate the incidence and prevalence of EoE in MPAACH, determine the features of EoE and other allergic comorbidity development, and elucidate the individual trajectories of allergic comorbidity development in children with AD that develop EoE.

## Methods

### Study Participants

The Mechanisms of Progression from Atopic Dermatitis to Asthma in Children (MPAACH) is a prospective early-life cohort of children with AD that have been detailed previously^4^. Briefly, children aged 0–2 years, had a gestation of ≥36 weeks, and either a diagnosis of AD (based on the Hanifin and Rajka Criteria for Atopic Dermatitis^7^) or the parent(s)/legal authorized representative indicates a positive response to each of the 3 questions from the Children’s Eczema Questionnaire^8^ were recruited between 2016 and 2024 and followed across 6 annual visits. This study was approved by the Institutional Review Board at Cincinnati Children’s Hospital Medical Center, and informed consent was provided by parents or guardians of participants.

### EoE Diagnosis

To determine EoE cases in MPAACH, we first used the electronic medical records to identify participants who had undergone endoscopy. Participants with ICD-10-CM code *K20*.*0* were flagged as potential EoE cases and were sent for clinical review. For those with available archived slides at our institution, distal esophageal biopsies from all endoscopies were retrieved and research eosinophil counts were performed. Peak eosinophil counts per high powered field were recorded for each endoscopy. Participants were classified as having EoE if they had at least 15 eosinophils per high-power field in at least one esophageal biopsy^9^.

### Skin Barrier Assessment

The children undergo assessment of the severity of their AD and the quality of their skin barrier function at each clinical exam. To assess AD severity, Scoring for Atopic Dermatitis (SCORAD)^10^ is performed. To determine the quality of the skin barrier, (TEWL) is determined at both lesional and never-lesional sites at each visit using the DermaLab TEWL probe (Cortex Technolog, Hadsund, Denmark).

As previously described, lesional skin was defined by the Hanifin and Rajka diagnostic criteria for atopic dermatitis^7^. Never-lesional skin was defined as at least 10 cm from current or historical lesional sites, and never-lesional sites had no history of lesions at that site according to parental report. Briefly, skin cells of lesional and never-lesional sites are sampled by placing the tape strip on the skin, gently massaging the tape for 15-20 seconds, removing the tape, and storing it in ice-cold buffer supplemented with 2% thio-glycerol (Promega, Madison, WI). This process is repeated to provide a total of 11 tape strips for each sampled skin site. To determine the level of *FLG* in the skin, we utilized RT-qPCR carried out with Taqman gene expression^4, 11^. The maximum gene expression value from either tape strip 8 or 9 from each subject, normalized to 18S levels, is used to denote *FLG* expression.

### Outcome Definitions

A positive skin prick test (SPT) is defined as a wheal measurement ≥3mm than the saline control to a food (n=15) or aeroallergen (n=11). Sensitization load was defined as the total number of sensitized allergens. Food allergy (FA) was defined based on parental report of a physician diagnosis of food allergy. Allergic rhinitis (AR) was defined as parental report of “a problem with sneezing or a runny or a blocked nose when he/she DID NOT have a cold or the flu,” and/or reported one or more allergy symptoms (itchy red eyes, runny nose, sneezing, itchy nose, congestion) and/or report of the child scratching or itching his/her eyes when he/she is in the same room with a cat, dog, disturbance of dust or near freshly cut grass, combined with at least one positive SPT result to an aeroallergen. Asthma was defined objectively by pulmonary function test results according to ATS criteria and healthcare utilization. Assessment of SPT, FA and AR are performed at all visits. Asthma is diagnosed at age 7-8.

### Statistical Analyses

Prior to the analysis, Shapiro-Wilk tests were performed to examine the distribution of the data for each continuous variable. Descriptive statistics are presented in median (25th-75th percentile) or frequency (%) due to the non-normal distributions. Comparisons between children with and without EoE were conducted using Wilcoxon rank-sum or Fisher’s exact tests for continuous and categorical variables, respectively. Statistical significance was defined as p-value<0.05. Statistical analyses were performed in R version 4.5.0 (R core team, 2024).

## Results

Of 700 MPAACH participants, 10 have EoE, and in all cases, EoE developed after AD onset. Two of the 10 subjects had EoE upon enrollment, while 8 developed EoE subsequent to enrolling in MPAACH. These 8 subjects developed EoE over the course of 1885 person-years, suggesting an incident rate of 42.5 EoE cases per 10,000 person-years. The cumulative incidence of EoE in MPAACH was 0.7% by age 2, 0.7% between ages 2 and 5, and 1.6% between ages 5 and 8 **(Figure 1A)**. The prevalence of EoE in MPAACH is 1.4% (95%CI 0.5-2.3%) and the median age at EoE diagnosis was 3.7 years (interquartile range (IQR): 1.8-6.5) **(Table 1)**. The prevalence of any allergic sensitization in MPAACH is 58.1% (95%CI 55.0-62.0%), and thus far, 23% (95%CI 19.6-25.8%) have developed FA, 41.3% (95%CI 34.5-48.0%) asthma, and 35.7% (95%CI 31.2-38.2%) AR; all higher than the general population^12^.

**Table 1.** Characteristics of MPAACH participants based on EoE Diagnosis.^1^.

|  | **AD children**<br>**with EoE**<br>**n = 10 (1.4%)** | **AD children**<br>**without EoE**<br>**n = 690 (98.6%)** | ***P*-value** |
| --- | --- | --- | --- |
| **Demographics** |  |  |  |
| Age at EoE diagnosis (y) | 3.7 (1.8-6.5) | - | - |
| Age at enrollment (y) | 2.3 (2.0-2.5) | 1.8 (1.0-2.4) | 0.07 |
| Male sex | 6 (60%) | 366 (53%) | 0.76 |
| Black race | 4 (40%) | 473 (69%) | 0.08 |
| Public insurance | 5 (50%) | 480 (70%) | 0.24 |
| **AD Severity** |  |  |  |
| SCORAD (score) | 20.1 (13.3-30.3) | 19.8 (12.3-30.7) | 0.92 |
| AD onset 0-3 months | 4 (40%) | 326 (47%) | 0.76 |
| **Skin barrier function** |  |  |  |
| **Lesional skin** |  |  |  |
| TEWL (g/m<sup>2</sup>/h) | 11.4 (7.5-25.1) | 12.5 (8.5-19.9) | 0.78 |
| Keratinocyte *FLG* expression | 0.47 (0.26-1.02) | 1.54 (0.38-4.10) | 0.068 |
| **Non-lesional skin** |  |  |  |
| TEWL (g/m<sup>2</sup>/h) | 11.5 (8.3-12.4) | 9.6 (7.2-13.4) | 0.80 |
| Keratinocyte *FLG* expression | 5.90 (1.34-8.34) | 2.58 (0.82-6.03) | 0.57 |
| **Sensitization** |  |  |  |
| Sensitization | 6 (60%) | 289 (42%) | 0.34 |
| Food sensitization | 6 (60%) | 196 (28%) | **0.039** |
| Aero sensitization | 4 (40%) | 191 (28%) | 0.48 |
| Sensitization load | 3 (0-4) | 0 (0-1) | **0.054** |
| **Development of comorbid conditions** |  |  |  |
| Asthma (N=190) | 4 (40%) | 70 (38%) | 0.42 |
| Food allergy | 8 (80%) | 154 (22%) | **<0.001** |
| Allergic rhinitis | 6 (70%) | 244 (35%) | 0.18 |

<sup>1</sup>Data are presented in median (25<sup>th</sup>-75<sup>th</sup>) or n (%). *P*-values were obtained using Wilcoxon rank-sum or Fisher's exact tests. AD: atopic dermatitis, EoE: eosinophilic esophagitis, *FLG*: filaggrin, SCORAD: scoring for atopic dermatitis, TEWL: trans-epidermal water loss.

**Figure 1.**
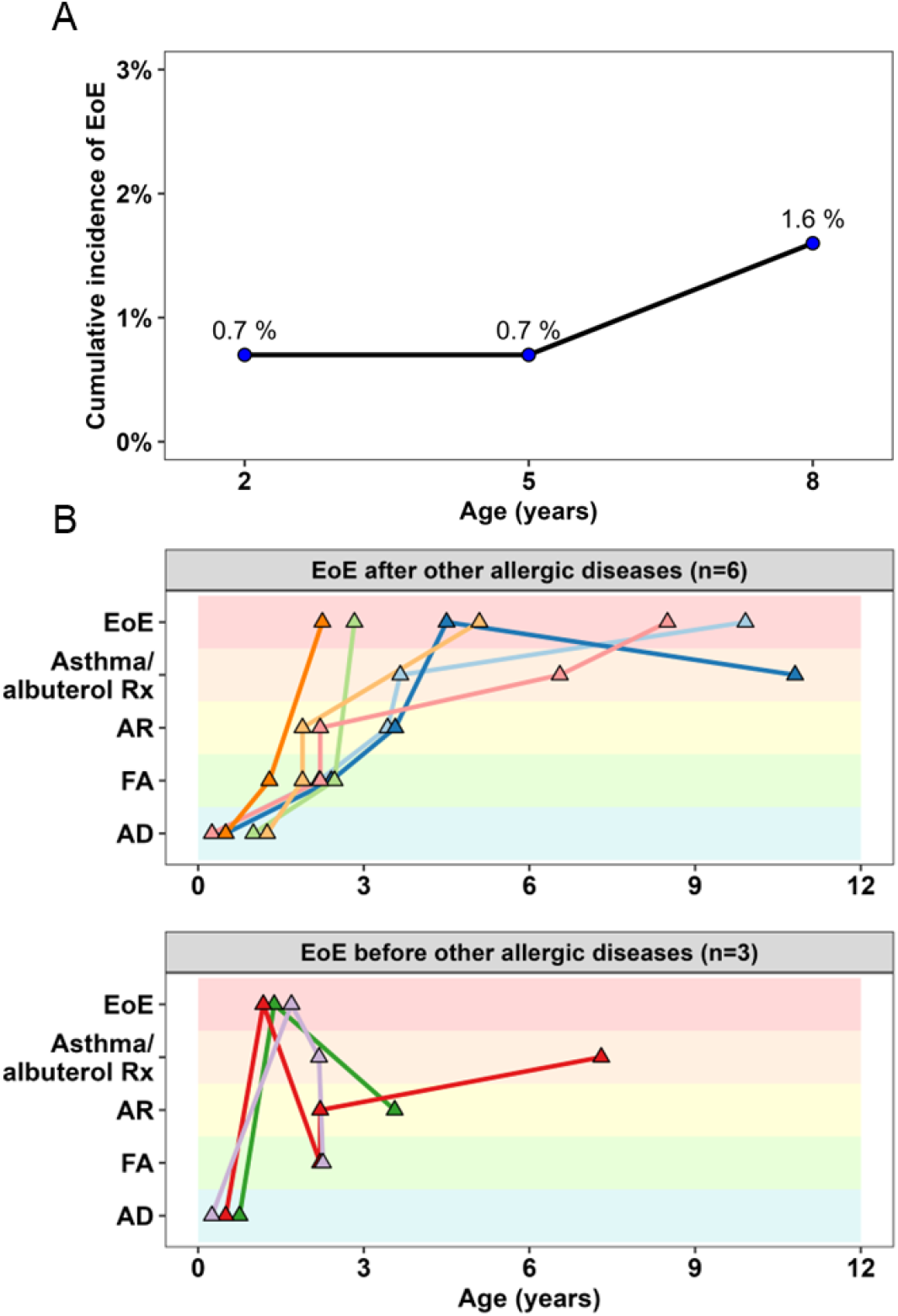
(A) Cumulative incidence of EoE at ages 2, 5, and 8 years and (B) the individual temporal sequences of allergic comorbidity onset. Cumulative incidence (CI) of EoE was calculated as the number of newly diagnosed EoE cases divided by the number of children at risk by age 2, between ages 2 and 5, and between ages 5 and 8. Children were considered at risk if they had not previously developed EoE and had reached the respective age. We then examined the sequence of allergic comorbidity onset among the 9 children with EoE who also had at least one allergic comorbidity besides AD. Triangle markers indicate the ages at which a specific allergic disease was diagnosed or reported for each participant.

There were no differences in demographics (race, sex, public insurance), AD severity (Scoring for Atopic Dermatitis (SCORAD)), trans-epidermal water loss (TEWL), keratinocyte filaggrin expression levels, or asthma prevalence between MPAACH children with or without EoE. Despite no difference in skin barrier quality or AD severity, children with EoE were more likely to have food sensitization as defined by at least one positive skin prick test to a food-allergen (60% vs. 28%, p=0.039), food allergy (70% vs. 13%, p<0.001), and showed a trend toward a higher sensitization load (3 (0-4) vs. 1 (0-1), p=0.054) compared to MPAACH children without EoE.

To date, 48.9% of children in MPAACH have developed at least one allergic comorbidity (FA, AR and/or asthma) by age 8 compared to 90% of the children with EoE (p=0.009), highlighting the strong association between EoE and other atopic diseases. To investigate temporal patterns, we examined the individual trajectories of atopic disease development including timing and sequence of allergic comorbidity onset among the 9 children with EoE who also had at least one allergic comorbidity in addition to AD **(Figure 1B)**. Of these, 6 (67%) children developed FA and/or AR prior to their EoE diagnosis, suggesting that EoE may emerge later in the allergic march. Notably, FA was present in all cases and AR in 4 of the 6 cases where EoE was preceded by another allergic condition. FA, AR, and/or asthma developed in 3 of 9 (33%) children after EoE onset **(Figure 1B)**.

## Discussion

Our data support that children with AD in the first 2 years of life have a higher risk for EoE. The prevalence of EoE in MPAACH is 1.4%, which is 10-fold higher than the general population^6^. Both AD and EoE are driven by a Th2-skewed immune response, characterized by elevated levels of interleukin-4 (IL-4), IL-5, and IL-13, which promote eosinophilia, IgE production, and barrier dysfunction^13^. In MPAACH, children with EoE had similar AD severity and skin barrier function compared to children with AD without EoE. However, children with EoE in MPAACH had significantly higher prevalences of food sensitization and food allergy, suggesting the esophageal epithelium may be the site of allergen penetration and immune activation, independent of the skin barrier. Indeed, growing evidence suggests that EoE involves epithelial dysfunction as the primary defect with secondary eosinophilic inflammation^14^.

Due to the shared pathophysiology of EoE with AD, FA, asthma and AR, it has been suggested that EoE is the 5^th^ member of the atopic march, with peak incidence after AD, FA, and asthma, and coinciding with AR^15^. Subjects in MPAACH did not exhibit this sequence of allergic disease development, likely due to study design. All MPAACH children have AD at age 0-2, asthma was not objectively diagnosed until age 7-8 years, and skin prick testing was performed at enrollment (age 0-2 years of age), potentially identifying AR earlier than in a clinical setting.

In conclusion, while most children with AD do not develop EoE, the significantly higher prevalence observed in the MPAACH cohort compared to the general population highlights the importance of early-life AD as an EoE risk factor. Given the shared Th2-driven immunopathology across AD, EoE, FA, asthma, and AR, our data support the inclusion of EoE as a fifth member of the atopic march. These findings underscore the need for heightened clinical awareness and proactive screening for EoE symptoms in children with early allergic manifestations.

## Data Availability

All data produced in the present study are available upon reasonable request to the authors.

## Abbreviations

AD: atopic dermatitis
AR: allergic rhinitis
EoE: eosinophilic esophagitis
FA: food allergy
MPAACH: Mechanisms of Progression of Atopic Dermatitis to Asthma in Children

## Acknowledgements

The authors would like to thank the MPAACH participants and their families.

## References

1. Hill DA, Spergel JM. The atopic march: Critical evidence and clinical relevance. Ann Allergy Asthma Immunol 2018; 120:131–7.

2. Davidson WF, Leung DYM, Beck LA, Berin CM, Boguniewicz M, Busse WW, et al. Report from the National Institute of Allergy and Infectious Diseases workshop on “Atopic dermatitis and the atopic march: Mechanisms and interventions”. J Allergy Clin Immunol 2019; 143:894–913.

3. Belgrave DC, Granell R, Simpson A, Guiver J, Bishop C, Buchan I, et al. Developmental profiles of eczema, wheeze, and rhinitis: two population-based birth cohort studies. PLoS Med 2014; 11:e1001748.

4. Biagini Myers JM, Sherenian MG, Baatyrbek Kyzy A, Alarcon R, An A, Flege Z, et al. Events in Normal Skin Promote Early-Life Atopic Dermatitis-The MPAACH Cohort. J Allergy Clin Immunol Pract 2020; 8:2285–93 e6.

5. Hill DA, Spergel JM. Is eosinophilic esophagitis a member of the atopic march? Ann Allergy Asthma Immunol 2018; 120:113–4.

6. Thel HL, Anderson C, Xue AZ, Jensen ET, Dellon ES. Prevalence and Costs of Eosinophilic Esophagitis in the United States. Clin Gastroenterol Hepatol 2025; 23:272–80 e8.

7. Hanifin JM, Rajka G. Diagnostic features of atopic dermatitis. Acta Derm Venereol 1980; 92 (suppl):44–7.

8. von Kobyletzki LB, Berner A, Carlstedt F, Hasselgren M, Bornehag CG, Svensson A. Validation of a parental questionnaire to identify atopic dermatitis in a population-based sample of children up to 2 years of age. Dermatology 2013; 226:222–6.

9. Dellon ES, Liacouras CA, Molina-Infante J, Furuta GT, Spergel JM, Zevit N, et al. Updated International Consensus Diagnostic Criteria for Eosinophilic Esophagitis: Proceedings of the AGREE Conference. Gastroenterology 2018; 155:1022–33 e10.

10. Severity scoring of atopic dermatitis: the SCORAD index. Consensus Report of the European Task Force on Atopic Dermatitis. Dermatology 1993; 186:23–31.

11. Stevens M, Gonzales T, Schauberger E, Baatyrbek Kyzy A, Andersen H, Spagna D, et al. Simultaneous skin biome and keratinocyte genomic capture reveals microbiome differences by depth of sampling J Allergy Clin Immunol 2020; In press.

12. Venter C, Palumbo MP, Sauder KA, Glueck DH, Liu AH, Yang IV, et al. Incidence and timing of offspring asthma, wheeze, allergic rhinitis, atopic dermatitis, and food allergy and association with maternal history of asthma and allergic rhinitis. World Allergy Organ J 2021; 14:100526.

13. Jaros J, Ahuja K, Lio P. Exploring the Link Between Atopic Dermatitis and Eosinophilic Esophagitis. J Clin Aesthet Dermatol 2025; 18:15–20.

14. Rochman M, Kellerman K, Jankowski MP, Rothenberg ME. The oesophagus as an immune organ. Nat Rev Gastroenterol Hepatol 2025; 22:657–67.

15. Hill DA, Grundmeier RW, Ramos M, Spergel JM. Eosinophilic Esophagitis Is a Late Manifestation of the Allergic March. J Allergy Clin Immunol Pract 2018; 6:1528–33.

